# COST AND UTILIZATION TRENDS OF PERCUTANEOUS KIDNEY BIOPSY IN KIDNEY TRANSPLANT PATIENTS: A 4-YEAR CLAIMS DATA ANALYSIS

**DOI:** 10.1101/2023.10.13.23295504

**Authors:** Adrian Vilalta

## Abstract

**Objective:** This study evaluated patterns of utilization and costs of percutaneous kidney biopsies in kidney transplant patients.

**Methods:** The IBM *Treatment Pathways* tool was used to interrogate cohorts selected from the IBM *MarketScan* dataset. Analyses were done for both adult and pediatric patients. Differences in biopsy paid amounts and utilization patterns were assessed for commercial payers, Medicare, and Medicaid. Claims data were collected for the years 2016 to 2019.

**Results:** A total of 15,337 records for percutaneous kidney biopsy events performed between 2016 and 2019 were identified in the database. Out of these, 840 corresponded to pediatric patients.

**Discussion:** Analysis showed that the paid amount for the procedure increased by 10% from 2016 to 2019 for commercial payers for adult patients, with US $4,137 being the median paid amount in 2019. Median paid amounts by both Medicare and Medicaid remained essentially flat for the same time at US $2,063 for Medicare and US $865 for Medicaid in 2019. Median cost of the procedure in pediatric patients increased 17% between 2016 and 2019 for commercial payers being US $6,068 in 2019. Medicaid payments for the pediatric patient population showed little change between 2016 and 2019, being US $1,666 in 2019.

## INTRODUCTION

Kidney transplantation is a lifesaving, last resort measure for people with end-stage kidney disease. It is estimated that close to 250 thousand people are alive today with a functioning kidney graft.^1^ However, kidney transplantation and graft health surveillance represent a significant financial burden. The estimated billed charges for a kidney transplant exceed US $440k and the average patient graft failure is predicted to incur additional lifetime medical costs of US $78,079.^2,3^ Percutaneous kidney biopsies are used routinely to both monitor the allograft’s health as well as to help diagnose any transplant complications such as rejection. Percutaneous kidney biopsy is commonly performed as an outpatient procedure under local anesthesia. Ultrasound is often used to guide the biopsy needle, determine kidney size, and detect the presence of cysts. Computed tomography imaging (CT) is used as an alternative for patients with small kidneys or high BMI.^4^ Although kidney biopsies are performed routinely, they present a risk of post-procedure complications ranging from pain at the biopsy site, to post-procedure hemorrhage, perinephric hematomas, and death.^5-7^

Despite being used routinely, there is a lack of current, real-world data on the financial cost of kidney biopsies, especially as it pertains to the kidney transplant population. Existing financial cost estimates are based on anecdotal, limited, outdated data sets or models.^8-10^ Importantly, actual payment data are required as bill charges overestimate actual payment for services.

The present study aimed to generate accurate, current, and complete data on the financial cost in US dollars of an outpatient kidney biopsy event by payer type for adults and pediatric kidney transplant patients.

## METHODS

Data presented here were obtained through a retrospective, observational claims-based analysis using a de-identified database to describe the patterns of utilization and episode costs of percutaneous kidney biopsies in kidney transplant patients.

### Patient Selection

Patients with evidence of a percutaneous kidney biopsy outpatient event and enrollment in either a commercial, Medicare, or Medicaid insurance plan were identified through the IBM® *MarketScan*® *Treatment Pathways*® database and analytical interface.^11^ *MarketScan*® collects all the coordination of benefits (COB) data thus generating total amount paid information, i.e., insurance, co-insurance, and patient pay. The event comprised all procedures performed the day of the kidney biopsy as defined by CPT 50200 in the claim. The cohort was further refined by selecting individuals with evidence of kidney transplantation. We also analyzed utilization and kidney biopsy costs in the pediatric population as defined by the American Academy of Pediatrics (21-years or younger).^12^ We obtained data for calendar years 2016 through 2019. During that period, the IBM® *MarketScan*® database contained the de-identified annual pooled healthcare data for over 48 million individuals with at least one year of data close to 200 million individuals insured commercially, or as part of the national distributed Supplemental Medicare and Medicaid programs throughout the entire United States. No IRB review was required given the nature of this study and the de-identified data source utilized.

### Study Measures

Evidence of transdermal kidney biopsy was established using Current Procedural Terminology (CPT) codes. The CPT code set is maintained by the American Medical Association.^13^ Diagnosis of kidney transplantation was established using International Classification of Diseases (ICD-9 and ICD-10) codes. ICD codes are created by the World Health Organization and contain codes for diseases, signs and symptoms, abnormal findings, complaints, social circumstances, and external causes of injury or diseases.^14^ Details on codes used in the present study can be found in **SUPPLEMENT 1**.

### Limitations

As with other claims-based studies, the findings may be impacted by the sensitivity of the claims data. Cohorts defined by this analysis do not consist of the complete universe of patients receiving the procedure. Only outpatient claims were included in the present analysis therefore costs of episodes systematically omit any inpatient services incurred through treatment of complications. Claims data may underestimate health care utilization among low-income individuals including Medicaid-insured and the non-insured population.

## STUDY RESULTS

Number of records identified for adult and pediatric patients from 2016 to 2019 are presented in **Figure 1**. Paid amounts per episode of care for adults are summarized in **Table 1**. Paid amounts for episode of care for pediatric patients (≤ 21 years) are presented in **Table 2**. We performed a proximate event analysis using the code for percutaneous kidney biopsy (CPT 50200) as the index code. This analysis identified procedures associated with CPT 50200 and their frequency the day of the kidney biopsy. The proximate event analysis was done to better understand the composition of procedures contributing to the total paid amount of the kidney biopsy episode of care. The list of the top 100 codes co-appearing in the kidney biopsy claims is provided in **SUPPLEMENT 2**.

**TABLE 1.**

| | Percutaneous Kidney Biopsy -<br>Paid Amounts per Episode of Care (US \$) Adults | |
| --- | --- | --- |
|  | Mean | Median |
| **2016** |  |  |
| **Commercial Insurance** | \$4,675 | \$3,778 |
| **Medicare Insurance** | \$3,956 | \$1,939 |
| **Medicaid Insurance** | \$1,645 | \$1,245 |
| **2017** |  |  |
| **Commercial Insurance** | \$4,985 | \$3,980 |
| **Medicare Insurance** | \$5,405 | \$1,951 |
| **Medicaid Insurance** | \$1,308 | \$706 |
| **2018** |  |  |
| **Commercial Insurance** | \$5,170 | \$4,180 |
| **Medicare Insurance** | \$6,436 | \$2,809 |
| **Medicaid Insurance** | \$1,504 | \$878 |
| **2019** |  |  |
| **Commercial Insurance** | \$5,404 | \$4,137 |
| **Medicare Insurance** | \$6,289 | \$2,063 |
| **Medicaid Insurance** | \$1,413 | \$865 |

**TABLE 2.**
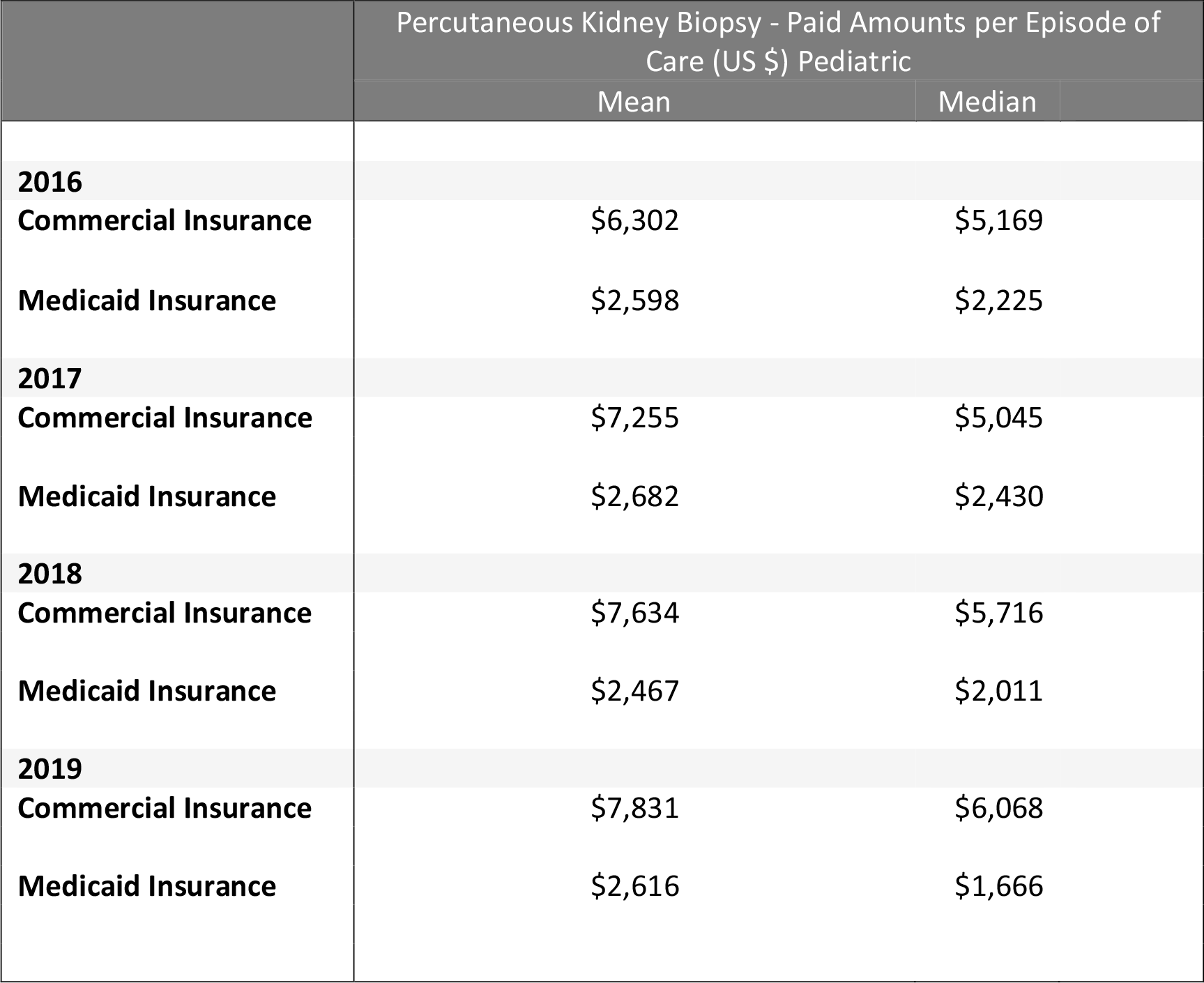

| | Percutaneous Kidney Biopsy - Paid Amounts per Episode of Care (US \$) Pediatric | |
| --- | --- | --- |
|  | Mean | Median |
| **2016** |  |  |
| **Commercial Insurance** | \$6,302 | \$5,169 |
| **Medicaid Insurance** | \$2,598 | \$2,225 |
| **2017** |  |  |
| **Commercial Insurance** | \$7,255 | \$5,045 |
| **Medicaid Insurance** | \$2,682 | \$2,430 |
| **2018** |  |  |
| **Commercial Insurance** | \$7,634 | \$5,716 |
| **Medicaid Insurance** | \$2,467 | \$2,011 |
| **2019** |  |  |
| **Commercial Insurance** | \$7,831 | \$6,068 |
| **Medicaid Insurance** | \$2,616 | \$1,666 |

**Figure 1.**
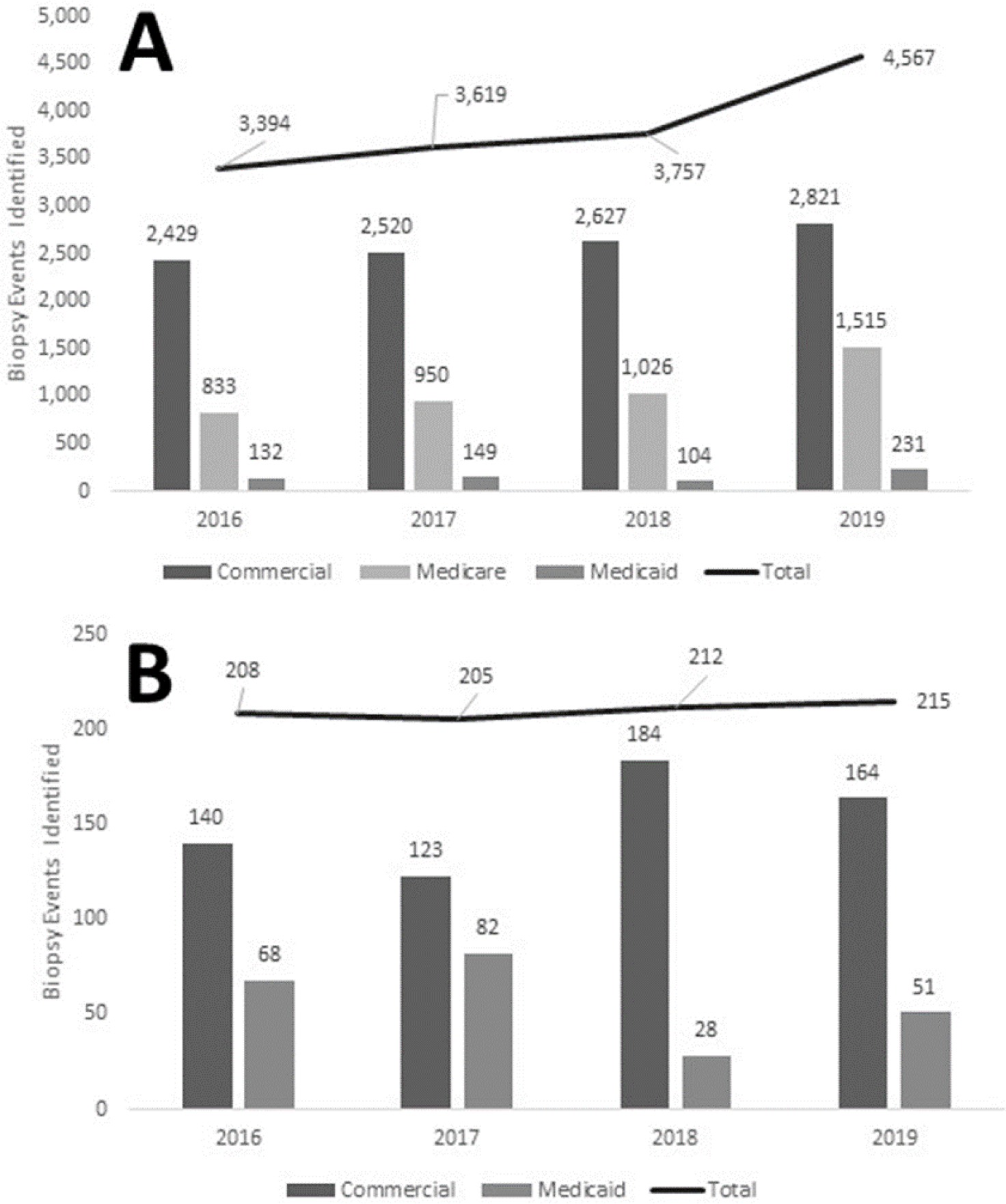
Percutaneous kidney biopsy records 2016 to 2019. (A) number of claims identified that met the inclusion criteria (i.e., CPT 50200 and evidence of kidney transplant) broken down by payer type. Total number of claims identified for each year is shown above the trendline. (B) number of claims identified for patient population 21 years of age or younger, broken down by payer type.

According to our analysis, the median paid amount for a percutaneous kidney biopsy event for an adult patient by commercial payers has increased steadily from 2016 ($3,778) to 2019 ($4,137) in kidney transplant patients. The increase is even higher (17%) for biopsies performed in the pediatric kidney transplant population, ranging from a median of $5,169 in 2016 to $6,068 in 2019. Remarkably, the paid amount for the procedure is close to 40 % higher in pediatric patients compared to the overall cohort, reflecting the complexities associated with performing the procedure in younger patients. While the paid amounts by commercial payers have increased, the paid amounts by both Medicare and Medicaid have remained relatively unchanged from 2016 to 2019. Our analysis indicated a median Medicare payment of $2,063 for the kidney biopsy event in 2019. According to Medicare, the national average total cost of the procedure in 2021 is $1,535, with $128 covering doctor’s fees, $1,407 facility fees, and the patient being responsible for $306.^15^ The higher paid amount revealed by our analysis reflects payment for additional services required the day of the biopsy event.

Our analysis also revealed a steady increase in the number of procedures performed year over year. The increase in the number of procedures from 2016 to 2019 ranges from about 16% if only commercially paid procedures are considered to close to 35% when procedures paid by Medicare and Medicaid are included. In the same period, the total number of kidney transplants reported in 2016 was 14,745 increasing to 18,018 in 2019, representing a 22 % increase in kidney transplantations.^16^ Although our data do not include every single transplant patient during the time window of the study, the increase in the identified kidney biopsy claims tracks with the increase in the number of kidney transplantations. However, estimates of the number of biopsies received by transplant patients and whether they have changed over time would require further analysis.

## CONCLUSIONS

Percutaneous kidney biopsy is a routine, recurrent procedure used to monitor the health of kidney allografts as well as to help diagnose potential complications. Spite its wide use, there is a paucity of recent, real-world data on paid amounts for the procedure, as well as the total cost of the episode of care. These data are essential to enable the development of clinico-economic models of clinical interventions that potential impact the number of subcutaneous kidney biopsies performed.

The present analysis bridged the data gap by providing paid amounts for the complete episode of care by payer type. The analysis also revealed significant differences in the cost of the procedure for pediatric kidney transplant patients compared to the kidney transplant population at large. In addition, the analysis showed trends in both the paid amount as well as the number of procedures performed. The cost and patterns of utilization established here provide a solid foundation for follow up analyses of the economic value of alternative allograft surveillance strategies.

## Data Availability

All data produced in the present study are available upon reasonable request to the authors.

---

**SUPPLEMENT 1.**
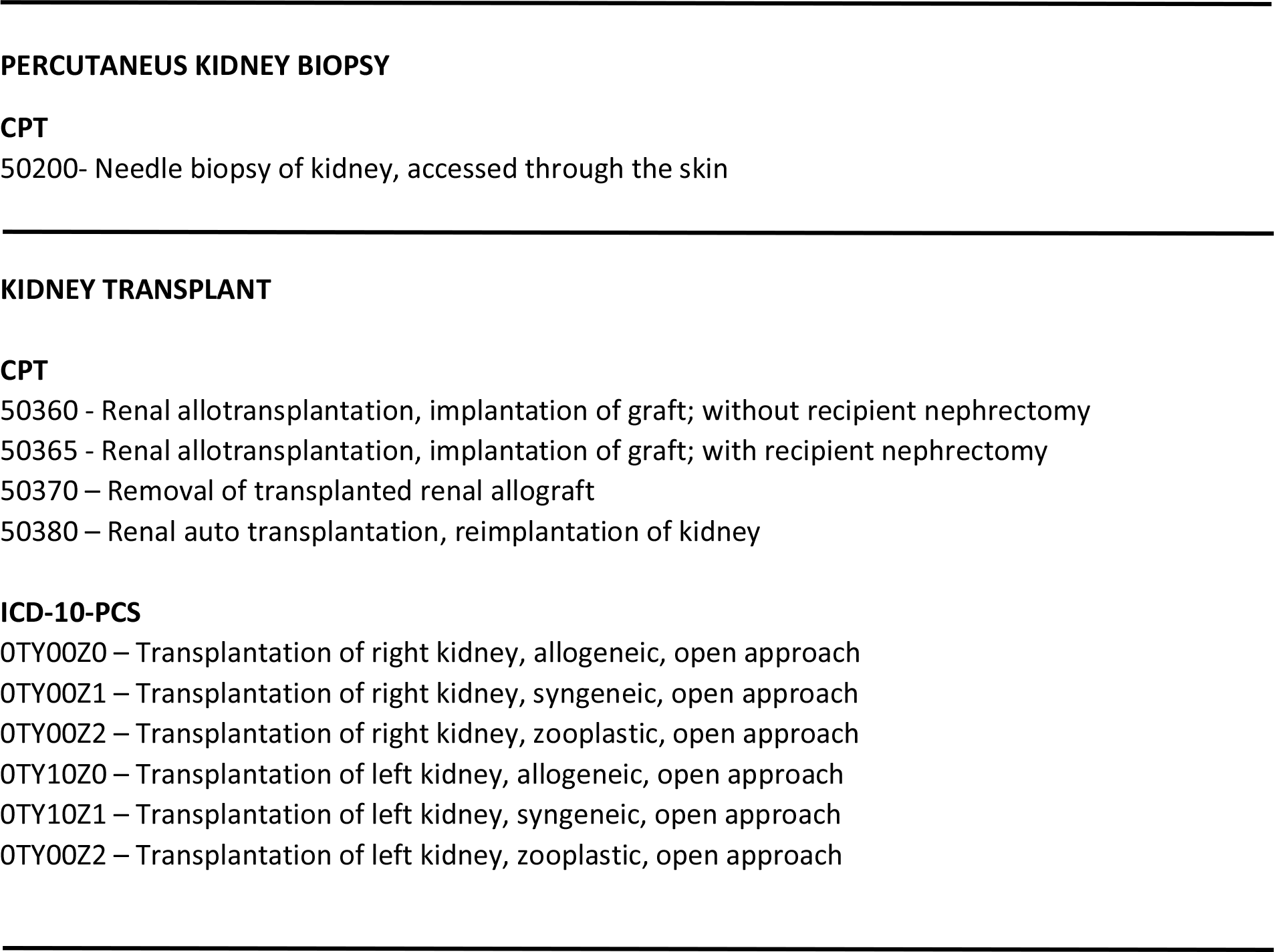
CODES USED TO IDENTIFY CLAIMS DATA.

**SUPPLEMENT 2.** MOST FREQUENT CODES CO-APPEARING IN OUTPATIENT KIDNEY BIOPSY CLAIMS.

| Code | Event Name | Frequency (%) |
| --- | --- | --- |
| 50200 | Renal biopsy; percutaneous, by trocar or needle | 100.0 |
| 88305 | Level IV - Surgical pathology, gross and microscopic examination<br>Abortion - spontaneous/missed Artery, biopsy Bone marrow, biopsy<br>Bone exostosis Brain/meninges, other than for tumor resection Breast,<br>biopsy, not requiring microscopic evaluation | 55.1 |
| 88313 | Special stain including interpretation and report; Group II, all other<br>(e.g., iron, trichrome), except stain for microorganisms, stains for<br>enzyme constituents, or immunocytochemistry and<br>immunohistochemistry | 47.1 |
| 88346 | Immunofluorescence, per specimen; initial single antibody stain<br>procedure | 36.0 |
| 88348 | Electron microscopy, diagnostic | 32.7 |
| 88350 | Immunofluorescence, per specimen; each additional single antibody<br>stain procedure (list separately in addition to code for primary<br>procedure) | 47.0 |
| 77012 | Computed tomography guidance for needle placement (e.g., biopsy,<br>aspiration, injection, localization device), radiological supervision and<br>interpretation | 33.8 |
| 85610 | Prothrombin time; | 23.9 |
| 76942 | Ultrasonic guidance for needle placement (e.g., biopsy, aspiration,<br>injection, localization device), imaging supervision and interpretation | 31.5 |
| 99152 | Moderate sedation services provided by the same physician or other<br>qualified health care professional performing the diagnostic or<br>therapeutic service that the sedation supports, requiring the presence<br>of an independent trained observer to assist in | 23.5 |
| J3010 | Injection, fentanyl citrate, 0.1 mg | 18.9 |
| J2250 | Injection, midazolam hydrochloride, per 1 mg | 18.7 |
| 36415 | Collection of venous blood by venipuncture | 13.3 |
| 88342 | Immunohistochemistry or immunocytochemistry, per specimen; initial<br>single antibody stain procedure | 18.2 |
| 85730 | Thromboplastin time, partial (PTT); plasma or whole blood | 13.8 |
| 85027 | Blood count; complete (CBC), automated (Hgb, HCT, RBC, WBC and<br>platelet count) | 12.5 |
| 85025 | Blood count; complete (CBC), automated (Hgb, HCT, RBC, WBC and<br>platelet count) and automated differential WBC count | 12.4 |
| 99153 | Moderate sedation services provided by the same physician or other<br>qualified health care professional performing the diagnostic or<br>therapeutic service that the sedation supports, requiring the presence<br>of an independent trained observer to assist in | 10.8 |
| 88341 | Immunohistochemistry or immunocytochemistry, per specimen; each additional single antibody stain procedure (List separately in addition to code for primary procedure) | 13.0 |
| 88329 | Pathology consultation during surgery; | 7.5 |
| 80048 | Basic metabolic panel (Calcium, total) This panel must include the following: Calcium, total (82310) Carbon dioxide (bicarbonate) (82374) Chloride (82435) Creatinine (82565) Glucose (82947) Potassium (84132) Sodium (84295) Urea nitrogen (BUN) (84520) | 7.7 |
| 88300 | Level I - Surgical pathology, gross examination only | 8.4 |
| 85014 | Blood count; hematocrit (HCT) | 5.1 |
| 85018 | Blood count; hemoglobin (Hgb) | 4.6 |
| 86900 | Blood typing, serologic; ABO | 4.0 |
| 86901 | Blood typing, serologic; Rh (D) | 4.0 |
| 86850 | Antibody screen, RBC, each serum technique | 3.9 |
| 81001 | Urinalysis, by dip stick or tablet reagent for bilirubin, glucose, hemoglobin, ketones, leukocytes, nitrite, pH, protein, specific gravity, urobilinogen, any number of these constituents; automated, with microscopy | 3.4 |
| 80053 | Comprehensive metabolic panel This panel must include the following: Albumin (82040) Bilirubin, total (82247) Calcium, total (82310) Carbon dioxide (bicarbonate) (82374) Chloride (82435) Creatinine (82565) Glucose (82947) Phosphatase, alkaline (84075) | 3.4 |
| J7030 | Infusion, normal saline solution, 1000 cc | 2.8 |
| J2001 | Injection, lidocaine HCl for intravenous infusion, 10 mg | 2.9 |
| 80197 | Tacrolimus | 3.0 |
| 85049 | Blood count; platelet, automated | 2.8 |
| J2405 | Injection, ondansetron hydrochloride, per 1 mg | 2.7 |
| 83735 | Magnesium | 2.8 |
| 84156 | Protein, total, except by refractometry; urine | 2.7 |
| G0378 | Hospital observation service, per hour | 2.8 |
| 82570 | Creatinine; other source | 2.6 |
| 87799 | Infectious agent detection by nucleic acid (DNA or RNA), not otherwise specified; quantification, each organism | 2.7 |
| 82962 | Glucose, blood by glucose monitoring device(s) cleared by the FDA specifically for home use | 2.8 |
| 80069 | Renal function panel This panel must include the following: Albumin (82040) Calcium, total (82310) Carbon dioxide (bicarbonate) (82374) Chloride (82435) Creatinine (82565) Glucose (82947) Phosphorus inorganic (phosphate) (84100) Potassium (84132) Sod | 2.5 |
| 84100 | Phosphorus inorganic (phosphate); | 2.3 |
| 860 | Anesthesia for extraperitoneal procedures in lower abdomen, including urinary tract; not otherwise specified | 2.5 |
| 88333 | Pathology consultation during surgery; cytologic examination (e.g., touch prep, squash prep), initial site | 2.9 |
| J7040 | Infusion, normal saline solution, sterile (500 ml = 1 unit) | 1.9 |
| 86832 | Antibody to human leukocyte antigens (HLA), solid phase assays (e.g., microspheres or beads, ELISA, Flow cytometry); high-definition qualitative panel for identification of antibody specificities (e.g., individual antigen per bead methodology), HLA Class | 1.9 |
| 82565 | Creatinine; blood | 1.9 |
| 86833 | Antibody to human leukocyte antigens (HLA), solid phase assays (e.g., microspheres or beads, ELISA, Flow cytometry); high-definition qualitative panel for identification of antibody specificities (e.g., individual antigen per bead methodology), HLA Class | 1.9 |
| 77013 | Computed tomography guidance for, and monitoring of, parenchymal tissue ablation | 2.6 |
| J2704 | Injection, propofol, 10 mg | 1.8 |
| 76776 | Ultrasound, transplanted kidney, real time, and duplex Doppler with image documentation | 2.7 |
| J3490 | Unclassified drugs | 2.2 |
| J0360 | Injection, hydralazine HCl, up to 20 mg | 1.5 |
| 88173 | Cytopathology, evaluation of fine needle aspirate; interpretation and report | 2.2 |
| 99219 | Initial observation care, per day, for the evaluation and management of a patient, which requires these 3 key components: A comprehensive history; A comprehensive examination; and medical decision making of moderate complexity. Counseling and/or coordination. | 1.3 |
| 862 | Anesthesia for extraperitoneal procedures in lower abdomen, including urinary tract; renal procedures, including upper one-third of ureter, or donor nephrectomy | 1.6 |
| 50593 | Ablation, renal tumor(s), unilateral, percutaneous, cryotherapy | 2.6 |
| 81025 | Urine pregnancy test, by visual color comparison methods | 1.5 |
| 84520 | Urea nitrogen; quantitative | 1.2 |
| 80061 | Lipid panel This panel must include the following: Cholesterol, serum, total (82465) Lipoprotein, direct measurement, high density cholesterol (HDL cholesterol) (83718) Triglycerides (84478) | 1.1 |
| 76775 | Ultrasound, retroperitoneal (e.g., renal, aorta, nodes), real time with image documentation; limited | 1.5 |
| 99214 | Office or other outpatient visit for the evaluation and management of an established patient, which requires at least 2 of these 3 key components: A detailed history; A detailed examination; Medical decision making of moderate complexity. Counseling | 1.1 |
| 83970 | Parathormone (parathyroid hormone) | 1.1 |
| 82947 | Glucose; quantitative, blood (except reagent strip) | 1.1 |
| 88172 | Cytopathology, evaluation of fine needle aspirate; immediate cytohistologic study to determine adequacy for diagnosis, first evaluation episode, each site | 1.3 |
| 99232 | Subsequent hospital care, per day, for the evaluation and management of a patient, which requires at least 2 of these 3 key components: An expanded problem focused interval history; An expanded problem focused examination; Medical decision making of | 1.3 |
| J1170 | Injection, hydromorphone, up to 4 mg | 1.0 |
| 99220 | Initial observation care, per day, for the evaluation and management of a patient, which requires these 3 key components: A comprehensive history; A comprehensive examination; and medical decision making of high complexity. Counseling and/or coordination. | 0.9 |
| 99233 | Subsequent hospital care, per day, for the evaluation and management of a patient, which requires at least 2 of these 3 key components: A detailed interval history; A detailed examination; Medical decision making of high complexity. Counseling and/or | 1.2 |
| 82306 | Vitamin D; 25 hydroxy, includes fraction(s), if performed | 1.0 |
| 84703 | Gonadotropin, chorionic (hCG); qualitative | 1.1 |
| 83036 | Hemoglobin; glycosylated (A1C) | 0.9 |
| J7050 | Infusion, normal saline solution, 250 cc | 0.9 |
| 88312 | Special stain including interpretation and report; Group I for microorganisms (e.g., acid fast, methenamine silver) | 1.1 |
| Q9967 | Low osmolar contrast material, 300-399 mg/ml iodine concentration, per ml | 0.8 |
| J1200 | Injection, diphenhydramine HCl, up to 50 mg | 0.8 |
| 81003 | Urinalysis, by dip stick or tablet reagent for bilirubin, glucose, hemoglobin, ketones, leukocytes, nitrite, pH, protein, specific gravity, urobilinogen, any number of these constituents; automated, without microscopy | 0.8 |
| 87086 | Culture, bacterial; quantitative colony count, urine | 0.8 |
| 87497 | Infectious agent detection by nucleic acid (DNA or RNA); cytomegalovirus, quantification | 0.8 |
| 99218 | Initial observation care, per day, for the evaluation and management of a patient which requires these 3 key components: A detailed or comprehensive history; A detailed or comprehensive examination; and Medical decision making that is straightforward | 0.7 |
| J2597 | Injection, desmopressin acetate, per 1 mcg | 0.7 |
| J0690 | Injection, cefazolin sodium, 500 mg | 0.7 |
| G9637 | At least two orders for the same high-risk medication | 0.7 |
| J7120 | Ringers lactate infusion, up to 1000 cc | 0.7 |
| 85576 | Platelet, aggregation (in vitro), each agent | 0.8 |
| 88307 | Level V - Surgical pathology, gross and microscopic examination<br>Adrenal, resection Bone - biopsy/curettings Bone fragment(s), pathologic fracture Brain, biopsy Brain/meninges, tumor resection Breast, excision of lesion, requiring microscopic evaluation. | 0.9 |
| 99215 | Office or other outpatient visit for the evaluation and management of an established patient, which requires at least 2 of these 3 key components: A comprehensive history; A comprehensive examination; Medical decision making of high complexity. Counseling. | 0.6 |
| 93010 | Electrocardiogram, routine ECG with at least 12 leads; interpretation and report only | 0.7 |
| J1100 | Injection, dexamethasone sodium phosphate, 1 mg | 0.6 |
| J2270 | Injection, morphine sulfate, up to 10 mg | 0.6 |
| 50592 | Ablation, 1 or more renal tumor(s), percutaneous, unilateral, radiofrequency | 1.3 |
| 80051 | Electrolyte panel This panel must include the following: Carbon dioxide (bicarbonate) (82374) Chloride (82435) Potassium (84132) Sodium (84295) | 0.6 |
| 76770 | Ultrasound, retroperitoneal (e.g., renal, aorta, nodes), real time with image documentation; complete | 0.9 |
| 88314 | Special stain including interpretation and report; histochemical stain on frozen tissue block (List separately in addition to code for primary procedure) | 0.6 |
| 96374 | Therapeutic, prophylactic, or diagnostic injection (specify substance or drug); intravenous push, single or initial substance/drug | 0.7 |
| C2618 | Probe/needle, cryoablation | 0.5 |
| 80180 | Mycophenolate (mycophenolic acid) | 0.5 |
| 99213 | Office or other outpatient visit for the evaluation and management of an established patient, which requires at least 2 of these 3 key components: An expanded problem focused history; An expanded problem focused examination; Medical decision-making. | 0.5 |
| 84550 | Uric acid; blood | 0.5 |
| 88334 | Pathology consultation during surgery; cytologic examination (eg, touch prep, squash prep), each additional site (List separately in addition to code for primary procedure) | 0.8 |
| 88331 | Pathology consultation during surgery; first tissue block, with frozen section(s), single specimen | 0.8 |

## REFERENCES

1. Scientific Registry of Transplant Recipients. (Accessed September 7, 2020, at https://www.srtr.org/.)

2. Bentley TS, Ortner NJ. Milliman Research Report U.S. organ and tissue transplants: Cost estimates, discussion, and emerging issues. 2020.

3. Sussell J, Silverstein AR, Goutam P, et al. The economic burden of kidney graft failure in the United States. Am J Transplant 2020;20:1323–33.

4. Korbet SM. Percutaneous renal biopsy. Semin Nephrol 2002;22:254–67.

5. Poggio ED, McClelland RL, Blank KN, et al. Systematic Review and Meta-Analysis of Native Kidney Biopsy Complications. Clin J Am Soc Nephrol 2020;15:1595–602.

6. Al Turk AA, Estiverne C, Agrawal PR, Michaud JM. Trends and outcomes of the use of percutaneous native kidney biopsy in the United States: 5-year data analysis of the Nationwide Inpatient Sample. Clin Kidney J 2018;11:330–6.

7. Aaltonen S, Finne P, Honkanen E. Outpatient Kidney Biopsy: A Single Center Experience and Review of Literature. Nephron 2020;144:14–20.

8. Dutta R, Okhunov Z, Vernez SL, et al. Cost Comparisons Between Different Techniques of Percutaneous Renal Biopsy for Small Renal Masses. J Endourol 2016;30 Suppl 1:S28–33.

9. Maripuri S, Penson DF, Ikizler TA, Cavanaugh KL. Outpatient versus inpatient observation after percutaneous native kidney biopsy: a cost minimization study. Am J Nephrol 2011;34:64–70.

10. McMahon GM, McGovern ME, Bijol V, et al. Development of an outpatient native kidney biopsy service in low-risk patients: a multidisciplinary approach. Am J Nephrol 2012;35:321–6.

11. IBM MarketScan Research Databases for Life Sciences Researchers. 2021. (Accessed 6/15/2021, at https://www.ibm.com/downloads/cas/OWZWJ0QO.)

12. Hardin AP, Hackell JM, Committee On P, Ambulatory M. Age Limit of Pediatrics. Pediatrics 2017;140.

13. The American Medical Association (AMA). CPT® (Current Procedural Terminology). (Accessed 6/2/2021, at https://www.ama-assn.org/amaone/cpt-current-procedural-terminology.)

14. The World Health Organization (WHO). International Statistical Classification of Diseases and Related Health Problems (ICD). (Accessed 6/2/2021, at https://www.who.int/standards/classifications/classification-of-diseases.)

15. Renal biopsy; percutaneous, by trocar or needle. Code: 50200. (Accessed 6/29/2021, at https://www.medicare.gov/procedure-price-lookup/cost/50200/.)

16. Organ Procurement and Transplantation Network. “National Data on the Candidate Waiting List, Organ Donation and Matching, and Transplantation.”. (Accessed 8/19/2020, at https://optn.transplant.hrsa.gov/data/.)

